# Distinct Spectral and Directional Thalamocortical Network Dynamics Define Focal Seizure Evolution

**DOI:** 10.64898/2026.02.03.26345480

**Authors:** Saarang Panchavati, Atsuro Daida, Sotaro Kanai, Shingo Oana, Hiroya Ono, Masaki Izumi, Kikuko Kaneko, Aria Fallah, Joe X Qiao, Noriko Salamon, Raman Sankar, Corey Arnold, William Speier, Hiroki Nariai

## Abstract

Neuromodulation targeting thalamic nuclei is increasingly used to treat drug-resistant focal epilepsy, yet human intracranial EEG studies describing how thalamocortical interactions evolve across seizures remain limited. We aimed to define frequency-specific thalamocortical network dynamics from seizure onset to termination, compare thalamocortical and cortico-cortical network activation, and test whether thalamic EEG features can classify seizure state to inform closed-loop or adaptive thalamic stimulation strategies. We retrospectively analyzed chronic stereo-EEG recordings from 19 patients with pediatric-onset, drug-resistant focal epilepsy (6 females; age at thalamic recording 1.0–28.1 years, median 16.9) with cortical and thalamic sampling. Sixty-six focal seizures were included. Spectral power, imaginary coherence, and spectral Granger causality were computed in non-overlapping two-second windows across slow (1–12 Hz), beta (13–30 Hz), and gamma (30–70 Hz) bands and compared with an interictal baseline. Random forest classifiers were trained using thalamic spectral power and thalamocortical connectivity features to distinguish ictal from non-ictal states using leave-one-patient-out cross-validation, with Shapley additive explanations used for feature attribution. Visual analysis identified thalamic ictal involvement at seizure onset in 82/101 thalamic contacts (81.2%), increasing to near-universal involvement by seizure termination, with onset-to-termination patterns dominated by low-voltage fast activity at onset and rhythmic spike or rhythmic slow-wave patterns at termination. The thalamus and cortical seizure onset zone exhibited broadband power increases at seizure onset that attenuated toward termination, while slow- and beta-band thalamocortical connectivity increased throughout seizures and peaked around the end-of-seizure epoch. Directed connectivity demonstrated bidirectional thalamocortical coupling, with slow-frequency thalamus-to-seizure onset zone outflow exceeding propagation-zone-to-seizure onset zone cortico-cortical outflow during both ictal and end-of-seizure epochs (anterior nucleus: p = 9.06 × 10L^3^ and p = 8.80 × 10L^3^; centromedian nucleus: p = 3.30 × 10L^3^ and p = 5.70 × 10L^3^). Seizure state was classifiable from thalamic spectral power and thalamocortical network features, achieving an area under the receiver operating characteristic curve of 0.825 ± 0.163 (anterior nucleus model) and 0.839 ± 0.149 (centromedian nucleus model), with thalamic broadband power plus slow-frequency thalamus-to-cortex outflow and beta-frequency cortex-to-thalamus inflow among the most informative features. Leveraging human intracranial EEG data, we define coordinated, frequency- and direction-specific thalamocortical and cortico-cortical network dynamics that evolve from seizure onset to termination. These findings establish a mechanistic basis and identify actionable thalamocortical EEG targets—particularly slow- and beta-band interactions—to inform individualized, adaptive, closed-loop neuromodulation aimed at optimizing seizure outcomes.

## Introduction

Epilepsy affects approximately 1% of the population, and nearly one-third of patients develop drug-resistant epilepsy.^1^ For individuals who are not candidates for curative resection, neuromodulation has emerged as an increasingly important therapeutic strategy. In particular, stimulation of thalamic nuclei, including the anterior and centromedian nuclei (AN and CM), is now widely used in both deep brain stimulation (DBS) and responsive neurostimulation (RNS) paradigms.^2–6^ Despite growing clinical adoption, the network mechanisms by which the thalamus participates in focal seizures—especially across seizure onset, propagation, and termination—remain understudied in human.

Prior intracranial electroencephalography (iEEG) studies have demonstrated thalamic involvement in both focal epilepsy across diverse seizure foci and generalized epilepsy, including tonic seizures and epileptic spasms.^7–14^ Visual analyses and spectral measures have shown that thalamic activity often emerges early during seizures and persists through their evolution.^7, 8^ In parallel, cortico-cortical interactions are well established contributors to seizure propagation, with increased synchronization observed across cortical networks from seizure onset to termination.^15–18^ However, these two lines of investigation have largely progressed independently. Consequently, it remains unclear how thalamocortical and cortico-cortical networks interact dynamically during focal seizures, and whether thalamocortical engagement represents a distinct component of the core ictal network rather than a secondary reflection of cortical spread.

Seizures are not broadband phenomena but are organized across specific frequency bands and directional pathways. Slow and beta frequency rhythms are particularly well suited to long-range synchronization and are known to be modulated by thalamocortical circuitry during physiological brain states as well as epileptic processes.^19–21^ Determining whether such frequency-specific, directional thalamocortical interactions are engaged during seizures is therefore critical, not only for clarifying seizure network organization but also for informing the design of adaptive or closed-loop neuromodulation strategies. Human stereo-electroencephalography (SEEG) uniquely enables this investigation by allowing simultaneous, nucleus-specific recordings from the thalamus and cortex with sufficient temporal resolution to characterize bidirectional, frequency-resolved network dynamics.

In this study, we leveraged chronic SEEG recordings from a pediatric-onset focal epilepsy cohort with simultaneous cortical and thalamic sampling to characterize thalamocortical network dynamics across seizure evolution. We analyzed spectral power, undirected connectivity, and directed connectivity from seizure onset through termination, with explicit comparison between thalamocortical and cortico-cortical interactions. In addition, we evaluated whether thalamic spectral and network features alone could reliably identify seizure state, thereby assessing their translational relevance for adaptive neuromodulation.

By integrating frequency-specific and directional network analyses across the full temporal course of seizures, this work aims to clarify the role of thalamocortical interactions within the broader ictal network and to establish mechanistic features of thalamic involvement that may be leveraged for individualized, closed-loop neuromodulation in focal epilepsy.

## Material and Methods

### Patient Cohort

Patients with pediatric-onset (before 18 years of age) epilepsy who were admitted to UCLA Mattel Children’s Hospital were recruited for this retrospective study. These patients underwent chronic SEEG implantation between November 2020 and July 2024, sampling an anticipated epileptic network that included the thalamus for possible future neuromodulation. Thalamic electrodes were targeted into the AN and/or CM. All patients had drug-resistant epilepsy, were suspected to have focal seizure onset, and were considered potential candidates for thalamic neuromodulation, including RNS and DBS. The justification for thalamic sampling using SEEG was carefully discussed within the context of clinical decision-making, and no additional intracranial electrodes beyond those deemed clinically necessary were placed for research purposes. Patients were excluded if their seizures consisted exclusively of brief (<10 s) or intermittent events (e.g., myoclonic seizures or epileptic spasms), which preclude analysis of temporal network dynamics.

### Standard Protocol Approvals

The research protocol was approved by the Institutional Review Board at the University of California, Los Angeles. Written informed consent was obtained for all surgical procedures, including SEEG implantation involving thalamic targets, as well as for the use of SEEG data for research purposes.

### Surgical Evaluation and Electrode Localization

SEEG implantation plans were discussed at UCLA’s multidisciplinary epilepsy surgery conference, consisting of epileptologists, neurosurgeons, neuroradiologists, and neuropsychologists. Implantation strategies were informed by seizure semiology, neurological examination, neuroimaging findings (MRI, PET, SPECT, and magnetoencephalography), neuropsychological evaluation, and scalp EEG, with primary emphasis on localization of the seizure onset zone (SOZ). Unilateral thalamic sampling was selected based on the suspected SOZ and its anatomical connectivity, whereas bilateral thalamic sampling was considered in cases with diffuse or poorly localized seizure onset.

Electrode trajectories were planned using BrainLab Elements software (Brainlab AG, Munich, Germany) based on gadolinium-enhanced and non-enhanced T1-weighted MRI sequences. Target thalamic nuclei were identified and manually outlined by an experienced neuroradiologist (NS) prior to trajectory planning.^10, 11^ Planned trajectories based on MP2RAGE sequences were coregistered to volumetric computed tomography for surgical guidance. Four-contact Spencer depth electrodes with 2.5 mm intercontact spacing were used for thalamic targets, while six- to twelve-contact electrodes with 5 mm spacing were used for cortical regions. Immediate postoperative computed tomography was performed to confirm electrode placement and exclude intracranial hemorrhage.

Cortical surface reconstruction was performed using preoperative MRI with FreeSurfer or Infant FreeSurfer for patients older than or younger than two years, respectively.^22, 23^ Electrode contact locations were verified using Brainstorm software,^24^ and thalamic electrode positions were further confirmed using automated thalamic segmentation with subsequent visual inspection.^25^

### EEG Acquisition and Preprocessing

SEEG recordings were obtained using a Nihon Kohden JE-120A amplifier (Irvine, CA, USA) at sampling frequencies of 200 Hz or 2,000 Hz, with a high-pass filter above 0.01 Hz, and stored in European Data Format. Data processing was performed using the MNE-Python library.^26^ SEEG signals were notch-filtered at 60 Hz and its harmonics up to half the sampling frequency to remove line noise. All recordings were subsequently downsampled to 200 Hz to ensure consistent sampling rates across seizures. A bipolar montage was applied to minimize the effects of volume conduction. Channels located in white matter, outside the brain, or containing prominent artifacts were excluded from analysis. Canonical frequency bands were defined as slow (1–12 Hz), beta (13–30 Hz), and gamma (30–70 Hz). Periods of interest were defined as follows: preictal (15 s before seizure onset), ictal (0–15 s after seizure onset), end-of-seizure (ES) (15 s before seizure termination), and postictal (15 s after seizure termination). For seizures shorter than 30 s, ictal and ES periods were divided at the seizure midpoint. An interictal baseline was defined as a one-minute segment occurring 20–19 minutes prior to seizure onset.

### Identification of Ictal Pattern

All seizures were reviewed to determine seizure onset and termination. Seizure onset was defined by the emergence of sustained rhythmic EEG patterns that were clearly distinct from the interictal baseline,^10, 11^ as identified by visual review by experienced clinical investigators (AD, SO, and HN). Seizure termination was defined as the disappearance of these sustained rhythmic EEG patterns and a return toward interictal activity. A maximum of three bipolar channels demonstrating the earliest and most prominent ictal involvement were designated as SOZ channels. Channels that exhibited ictal changes within 10 seconds of seizure onset were labeled as propagation channels, and all remaining channels were annotated as other channels.

Thalamic ictal seizure patterns were independently assessed by two board-certified epileptologists (AD and SK). For each seizure, the reviewers annotated (i) involvement of the thalamus at seizure onset, (ii) thalamic onset EEG pattern, (iii) involvement of the thalamus at seizure offset, and (iv) thalamic offset EEG pattern. Thalamic EEG patterns were categorized based on previously established classifications^8^ into the following categories: none, low-voltage fast activity (LVFA), rhythmic spike, spike-and-wave or rhythmic slow-wave activity (RS/RSw), and slow wave without rhythmicity. Multiple ictal EEG patterns could be present simultaneously within the thalamus during a given seizure.

When LVFA co-occurred with any other ictal EEG pattern, LVFA was designated as the primary pattern. RS/RSw was annotated when spike-and-wave or rhythmic slow-wave activity was the dominant pattern in the absence of clear fast activity. The slow-wave category was selected when an increase in slow-frequency power without rhythmicity was observed and neither LVFA nor RS/RSw criteria were met. In cases of disagreement between AD and SK, final classification was determined by consensus discussion with a third reviewer (HN).

After completion of all ratings, interrater reliability between the two primary reviewers was assessed using Cohen’s kappa for the following measures: (i) presence of thalamic involvement at seizure onset, (ii) thalamic onset EEG pattern, (iii) presence of thalamic involvement at seizure offset, and (iv) thalamic offset EEG pattern.

For post hoc analyses, each seizure was further classified according to seizure focus, ictal onset pattern, and presence of diffuse involvement at onset. Seizure focus was defined by lobar localization of the SOZ, including frontal, parietal, temporal, occipital, and limbic regions, with limbic regions comprising the hippocampus, parahippocampus, amygdala, cingulate gyrus, and insula. Ictal onset patterns were classified as LVFA, RS/RSw, or other, with the latter category including seizures characterized by slow-wave activity or no clear ictal pattern at onset. Diffuse involvement at seizure onset was defined as early ictal involvement (SOZ and propagation channels) spanning bilateral hemispheres or three ipsilateral systems (e.g., left frontal, temporal, and limbic regions).

### EEG Analysis

For each seizure, we computed spectral power, imaginary coherence, and spectral Granger causality during the predefined periods of interest and interictal baseline periods. To characterize temporal evolution, each metric was calculated in non-overlapping two-second windows spanning from two minutes before seizure onset to two minutes after seizure termination. To mitigate intersubject variability, spectral power and connectivity measures were expressed as either percent change or log-ratio relative to the interictal baseline, as appropriate. Analyses focused on the AN and CM ipsilateral to the SOZ (Ip AN and Ip CM). Metrics were first computed at the level of individual SEEG channels and subsequently averaged within anatomically defined regions to yield a single measure per region per time window.

### Spectral Analysis

Power spectral density (PSD) was computed for each period of interest, time window, channel, and frequency band. Canonical frequency bands were defined a priori as slow (1–12 Hz), beta (13–30 Hz), and gamma (30–70 Hz). Spectral power values were averaged across time windows corresponding to each seizure phase (interictal, preictal, ictal, ES, and postictal).

### Imaginary Coherence (iCoh)

Coherence is an undirected measure of synchronization and correlation between two time series signals at different frequencies.^27^ Values of coherence range between 0 and 1, where 1 represents the highest degree of synchrony between two signals. Coherence has widely been used in EEG data analysis to understand functional connectivity between different brain regions.^11, 27^ Traditional coherence measures are susceptible to volume conduction effects, which can artificially inflate connectivity estimates. The iCoh isolates the imaginary component of the cross-spectrum and thus captures non-zero phase-lagged interactions between signals. Imaginary coherence was computed using the MNE-connectivity Python package.^26^ For each seizure, iCoh was calculated between all channel pairs and subsequently grouped according to anatomical location, including SOZ, Ip AN, Ip CM, propagation channels, and other channels. iCoh values were then averaged within each interaction type to yield robust region-to-region connectivity measures across time and frequency bands.

### Spectral Granger Causality (GCA)

While imaginary coherence provides an undirected measure of functional connectivity, it does not capture the direction of information flow. In the time domain, Granger causality analysis (GCA) quantifies whether past values of one signal improve prediction of another signal beyond its own history, and it has been widely applied in neuroscience to characterize directed measures of connectivity, including epilepsy research.^11, 12, 28, 29^ The spectral extension enables frequency-domain estimation of directional interactions. Spectral GCA was computed using the spectral-connectivity Python package.^30^ Directed connectivity was defined as thalamic inflow (SOZ → thalamic nucleus) and thalamic outflow (thalamic nucleus → SOZ), allowing assessment of bidirectional thalamocortical influence across seizure evolution and frequency bands.

## Random Forest Classification of Seizure State

To evaluate the translational utility of thalamic signals for real-time seizure detection, we used random forest classifiers to distinguish ictal from non-ictal states. Random forests were chosen for their robustness to nonlinear feature interactions, correlated predictors, and moderate sample sizes, as well as their strong empirical performance in biomedical classification tasks.^31^ EEG data were epoched from seizure onset to termination, with an additional one-minute preictal and postictal window. Each seizure was segmented into non-overlapping two-second windows. Separate random forest classifiers were trained for AN and CM features. Feature sets included thalamic spectral power, iCoh, and spectral GCA between the thalamus and SOZ. Each model classified whether a given time window corresponded to an ictal or non-ictal state.

Model performance was evaluated using leave-one-patient-out cross-validation, and performance was quantified using the area under the receiver operating characteristic curve (AUROC) for each seizure when it appeared in the test set. To improve interpretability, SHAP (SHapley Additive exPlanations)^32^ were computed across all cross-validation folds and averaged to estimate the magnitude and direction of feature importance. Ablation analyses were performed by systematically removing feature categories (e.g., spectral power only, connectivity only, or specific frequency bands) to assess their relative contributions to classification performance.

## Statistical Analysis

For group-level comparisons, two-tailed Wilcoxon signed-rank tests were used to assess whether spectral power, iCoh, or spectral GCA values differed significantly from the interictal baseline. For comparisons between non-overlapping network groups (e.g., thalamocortical vs cortico-cortical interactions), Mann–Whitney U tests were applied. Data were grouped by seizure and averaged prior to statistical testing. All p values were corrected for multiple comparisons using Bonferroni adjustment, accounting for the number of frequency bands, interaction types, and time windows tested.

## Data Availability

The EEG data and analysis code supporting the findings of this study will be made publicly available upon publication in a peer-reviewed journal, with EEG datasets hosted on OpenNeuro and all analysis code accessible via GitHub, to promote transparency, reproducibility, and open science.

## Results

### Patient demographics

A total of 21 patients were initially identified for this study. Two patients were excluded because their recorded events consisted exclusively of epileptic spasms or focal seizures followed by epileptic spasms, with individual ictal events lasting less than 5 seconds, precluding analysis of seizure dynamics. Consequently, 19 patients (6 females) were included in the final cohort (**Table 1**). The median age at thalamic recording was 16.9 years (range, 1.0–28.1 years). In total, 1,599 SEEG contacts were implanted, including 101 thalamic contacts (51 in AN and 50 in CM) (**Figure 1**). Each patient had a median of 83 contacts implanted (range, 38–148). Unilateral thalamic sampling was performed in 18 patients, while one patient underwent bilateral thalamic sampling. A total of 66 focal seizures were analyzed, with a median of 3 seizures per patient (range, 1–8).

| Age category* (yr) | Sex | Epilepsy type/ Etiology | AEDs during SEEG | MRI/PET hypometabolism | Prior Surgery | Onset | Non-thalamic coverage | Thalamic Coverage | Seizure Onset Pattern | Seizure End Pattern | Semiology | Outcomes (intervention) |
| --- | --- | --- | --- | --- | --- | --- | --- | --- | --- | --- | --- | --- |
| 0-2 | M | LGS /Genetic-Structural(TSC) | CBD, CLB, FEB, LEV | Bilateral cortical & subcortical Tub, subependymal nodules/L F,T,P | L Tub laser ablation, CC, L T lobectomy, & L O Tub resection | R F: 1 | R: F, P, T<br>L: F | RAN, RCM | RAN:none, RCM: LVFA | RAN:none, RCM: LVFA | FMS | No surgery |
| 6-12 | M | Focal /Structural (R ethmoid encephalocele) | LCM,LEV, BTM | R F gliosis / R F | R F pole resection | R P;3 | R: F, T, P, LIM<br>L: F, T, P, LIM | RAN, RCM | RAN:LVFA/Slow, RCM: LVFA | RAN:RS/RSw, slow RCM: RS/RSw | FNMS | RNS (Incl. RCM) |
| 18-29 | F | Rasumussen syndrome / Structural | BTM, CBD,CLB, ESL, FBM, ZNS | L putamen FLAIR hyperintensity; L F, T, BG atrophy( & L lateral ventricle dilation)/ L F | VNS | L T:3 | R: T<br>L : F, P, T | LAN, LCM | LAN: LVFA, none; LCM: LVFA, none | LAN: RS/RSw, LCM: RS/RSw | FMS | RNS (Incl. LCM) |
| 18-29 | F | Focal / Structural (FCD) | BVT, CLB, CNB | L T,P,O gliosis/ L TPO | L T resection, VNS | L LIM: 2 | L: F, P, O, T, LIM | LAN, LCM | LAN: LVFA | LAN: RS/RSw | FMS<br>FNMS | RNS (Incl. LAN) |
| 0-2 | M | Focal / Structural ( FCD/NRPL3) | OXC, VGB | None/ R T | none | R T:3 | R: F, T, LIM, L :F | RCM | RCM: LVFA, RS/RSw | RCM: RS/RSw | FNMS | Laser ablation for FCD |
| 0-2 | M | FS+ post IESS/Genetic-structural (TSC) | CLB, LEV, PER, VGB | Bilateral cortical & subcortical Tub, subependymal nodules/Bilateral F,T,P,O | none | R F: 3 | R: F, T<br>L: O, LIM | RAN,RCM | RAN: LVFA, none RCM:LVFA, none | RAN: RS/RSw RCM: RS/RSw | FNMS | Right tuberectomy |
| 13-17 | F | Focal / Structural (FCD) | LTG, OXC | None (lack of left brain) /L F | L subtotal hemispherectomy; L sensorimotor resection | L F: 2, L P: 3 | L: F, P, LIM | LCM | LCM: RS/RSw, LVFA | LCM: RS/RSw | FMS, FBTCs | RNS (Incl. LCM) |
| 13-17 | F | LGS /Genetic-Structural(Bandheterotopia) | FFA, CNB, CLB, RUF | Posterior dominant bandheterotopia/ NL | VNS | L O: 3 | R: O, L: O | RAN,RCM | RAN: LVFA, RS/RSw, none RCM: LVFA | RAN: RS/RSw RCM: RS/RSw | FMS | RNS (LCM & RPUL) |
| 3-5 | M | Focal/Structual (perinatal ischemia) | CLB,LTG, PER | RF Porencephalic syst / L T | none | L O: 2 | R: F, P, T, O<br>L: P, O, LIM | LCM | LCM: RS/RSw, none | LCM: slow, none | FNMS | RNS (Incl. LPUL) |
| 6-12 | M | Focal/Structual (periventricular gray matter heterotopia) | LTG | LF Gray matter heterotopia/NL | none | L F:3 | R: F, LIM<br>L: F, P, T, LIM | LAN, LCM | LAN: RS/RSw LCM: RS/RSw | LAN: RS/RSw, slow, none LCM: RS/RSw, slow, none | FNMS | RNS (Incl. LCM) |
| 13-17 | F | Focal/Genetic-structural (TSC) | CNB, LTG, ZNS | Subcortical Tub L F & R F,T,P, subependymal nodules/L F | none | L F:3 | R: F, T, P<br>L: F, T, LIM | LAN, LCM | LAN: LVFA, NA<br>LCM: None | LAN: LVFA, slow, none<br>LCM: LVFA, slow, none | FNMS | Left tuberectomy |
| 18-29 | M | Focal/Unknown | CLB, LTG | R T FLAIR hyperintensity(or none?)/ NL | none | R F:3 | R: F, T, LIM<br>L: F, T, LIM | RAN, RCM | RAN: RS/RSw<br>RCM: RS/RSw | RAN: RS/RSw<br>RCM: RS/RSw | FNMS, FBTCS | No surgery |
| 13-17 | M | Focal/Structural (FCD) | CBD, CBZ, CLB, LCM, Ketogenic Diet | L insula FCD / L insula | none | L LIM: 3 | L: F, P, T, LIM | LAN, LCM | LAN: LVFA, NA<br>LCM: LVFA, NA | LAN: RS/RSw<br>LCM: RS/RSw | FMS, FBTCS | RNS (Incl. LAN) |
| 18-29 | M | Focal/Unknown | BTM, CNB, LCM | NL / NL | R T lobectomy | R T:3 | R: F, P, T, O, LIM | RAN, RCM | RAN: RS/RSw<br>RCM: RS/RSw | RAN: RS/RSw<br>RCM: RS/RSw | FNMS | RNS (Incl. RAN) |
| 18-29 | M | Focal / Structural (Encephalitis & FCD) | CNB, LCM, LEV | L T FLAIR hypterintensity / NL | VNS | R T: 3<br>R P:1<br>L T:4 | R: F, T, P, LIM<br>L: F, T, P, LIM | RAN, LAN | LAN: LVFA, RS/RSw, Slow, none<br>RAN: LVFA, RS/RSw, none | LAN: RS/RSw, slow, none<br>RAN: RS/RSw, slow, none | FNMS | RNS (bil. AN) |
| 13-17 | M | Focal / Unknown | CBD, OXC, CLB | NL / L T,P | none | L P:3<br>L LIM:1 | R:F, P, O, T, LIM<br>L: F, P,O, T, LIM | LAN, LCM | LAN: LVFA, RS/RSw<br>LCM: LVFA | LAN: RS/RSw<br>LVFA: RS/RSw | FNMS, FBTCS | RNS (Incl. RCM) |
| 18-29 | F | Focal / Unknown | LTG, LEV | NL / L T | VNS | L LIM: 4 | L: F,T,LIM,BG | LAN, LCM | LAN: RS/RSw<br>LCM: RS/RSw | LAN: RS/RSw<br>LCM: RS/RSw, slow | FMS, FNMS | RNS |
| 18-29 | M | Focal / Structural (FCD) | CNB, OXC, CZP | L P & periventricular white matter T2 FLAIR hyperintensity | L hemicraniotomy | R P: 3<br>(L P: 2) | R:T,P, LIM<br>L:T,P, LIM | LAN, LCM | LAN: LVFA, RS/RSw, none<br>LCM: LVFA, RS/RSw, none | LAN: LVFA, RS/RSw<br>LCM: LVFA, RS/RSw | FNMS, FBTCS | RNS (Incl. LCM) |
| 18-29 | M | Focal / Unknown | LEV, LTG, RUF, CLB | NL/NL | None | R F: 4 | R: F, T, LIM, P<br>L :F, T, LIM | RAN, RCM | RAN: LVFA<br>RCM: LVFA | RAN: RS/RSw<br>RCM: RS/RSw | FMS | RNS (Incl. RCM) |

AN, anterior nucleus of the thalamus; BTM, brivaracetam; CBD, cannabidiol; CBZ, carbamazepine; CLB, clobazam; CM, centromedian nucleus of the thalamus; CNB, cenobamate; CZP, clorazepate; ESL, eslicarbazepine; F, frontal; FBM, felbamate; FBTCS, focal to bilateral; FFA, fenfluramine; FMS, focal motor seizure; FNMS, focal non-motor seizure; IEES, Infantile Epileptic Spasms Syndrome; LEV, levetiracetam; LGS, Lennox-Gastaut Syndrome; LTG, lamotrigine; L left; LCM, lasocamide; LIM, limbic system; LVFA, low voltage fast activity; NL, non-lesion; O, occipital; OXC, oxcarbazepine; P, parietal; PER, perampanel; PUL, pulvinar; T, temporal; TSC, Tuberous Sclerosis Complex ; Tub, tuber; R; right; RS/RSw, rhythmic spike, spike-and-wave, or slow wave; RNS, responsive neurostimulation; RUF, rufinamide; Slow, slow wave without rhythmicity; VGB, vigabatrine; VNS
\*Age categories were defined as inclusive intervals based on chronological age at evaluation, with the “0–2 years” category encompassing ages from birth through 2 years 11 months.

**Figure 1.**
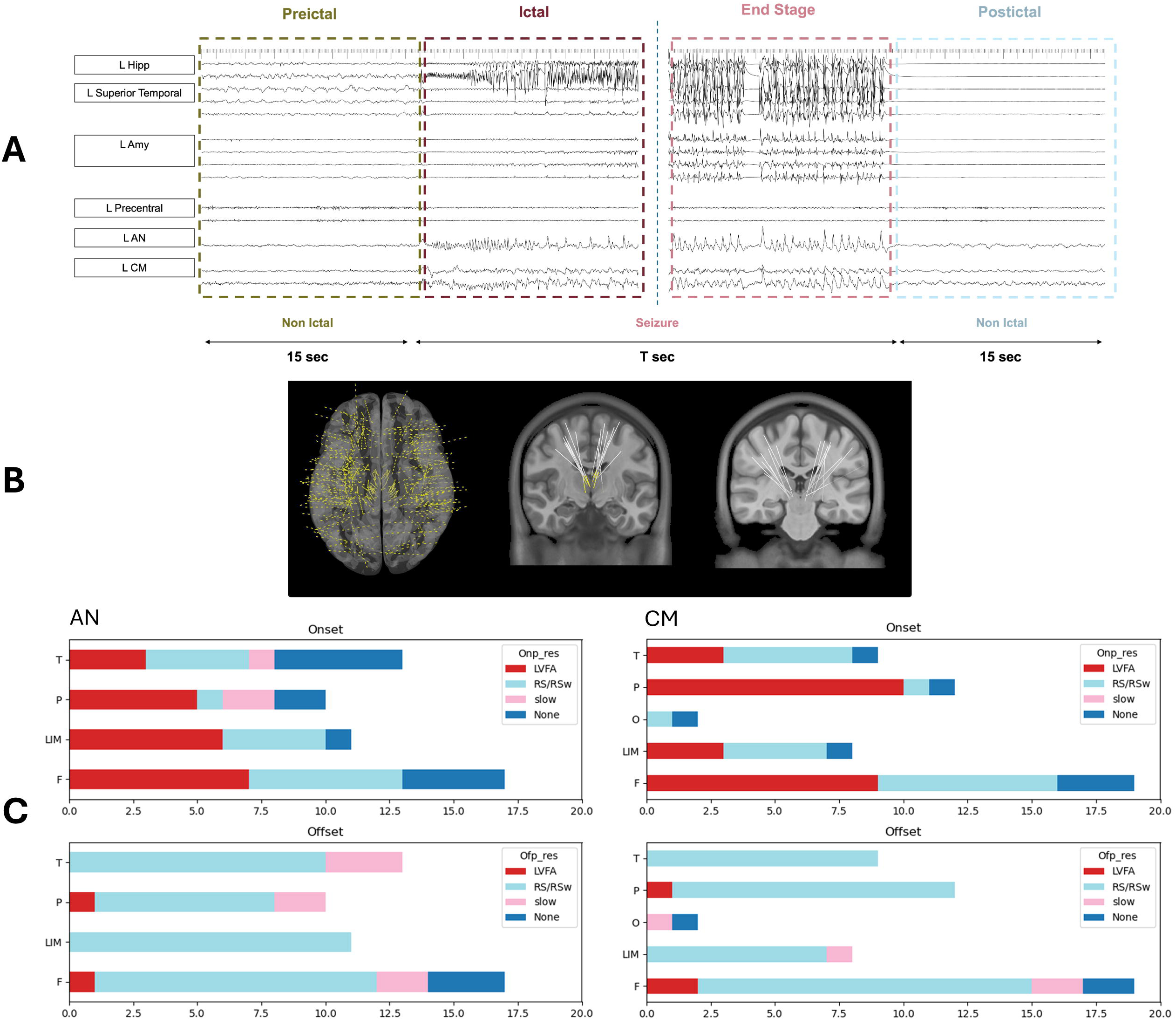
Overview of approach and visual characterization of thalamic ictal patterns. (A) Representative SEEG seizure example illustrating the analysis epochs: preictal (15 s), ictal (seizure duration, T s), end-of-seizure (15 s preceding termination), and postictal (15 s). Traces are shown from representative cortical regions and ipsilateral thalamic contacts (AN and CM). Vertical dashed lines denote seizure onset and seizure termination. (B) Example electrode localization demonstrating widespread cortical sampling and thalamic depth electrode trajectories targeting AN and CM (all subjects combined). (C) Distribution of visually classified thalamic ictal patterns at seizure onset (top) and seizure termination (bottom), shown separately for AN contacts (left) and CM contacts (right), stratified by SOZ focus (F, frontal; LIM, limbic; T, temporal; P, parietal; O, occipital). Patterns are categorized as LVFA, RS/RSw, slow, or none.

### Visual analysis

Visual analysis revealed that most thalamic contacts (82/101, 81.2%) exhibited ictal changes at seizure onset. Within the AN (n = 51), ictal activity was observed at seizure onset in 39 contacts (76.4%), comprising LVFA in 21 (41.2%), RS/RSw in 15 (29.4%), and slow-wave activity in 3 (5.9%). At seizure termination, ictal changes were present in 48 AN contacts (94.1%), with LVFA in 2 (3.9%), RS/RSw in 39 (76.5%), and slow-wave activity in 7 (13.7%). Within the CM (n = 50), ictal changes at seizure onset were observed in 43 contacts (86.0%), consisting of LVFA in 25 (50.0%) and RS/RSw in 18 (36.0%). At seizure termination, ictal activity was present in 47 CM contacts (94.0%), with LVFA in 3 (6.0%), RS/RSw in 40 (80.0%), and slow-wave activity in 4 (8.0%) (**Figure 1**).

Overall, thalamic ictal patterns evolved from a predominance of LVFA at seizure onset toward RS/RSw patterns at seizure termination across both thalamic nuclei. Interrater agreement assessed using Cohen’s kappa demonstrated substantial agreement for annotation of ictal onset involvement (κ = 0.78) and onset pattern classification (κ = 0.82), and moderate agreement for ictal offset involvement (κ = 0.63) and ictal offset pattern classification (κ = 0.57).

### Frequency-specific thalamic and SOZ spectral power changes across seizure periods

Spectral power changes in the AN, CM, and SOZ were compared with the interictal baseline across predefined seizure periods. Time-domain analyses demonstrated broadband power increases during seizures, evident during both the ictal and ES periods, followed by a rapid decrease after seizure termination in both the thalamus and SOZ (**Figure 2A,B**). Within the thalamus, slow- and beta-band activity predominated toward seizure termination in both nuclei (**Figure 2C**). Significant slow-band power increases were observed in thalamic channels during the ictal and ES periods, with the largest effects at ES (**Figure 2D**; AN: p = 6.18 × 10LL; CM: p = 5.58 × 10LL). Beta- and gamma-band power also increased at seizure onset, with beta-band activity persisting through ES (**Figure 2B**). In the SOZ, beta-and gamma-band power showed significant increases at seizure onset and ES, whereas slow-band power increased primarily during ES (**Figure 2D**). Notably, beta- and gamma-band activity in the SOZ decreased during the postictal period, returning toward baseline levels.

**Figure 2.**
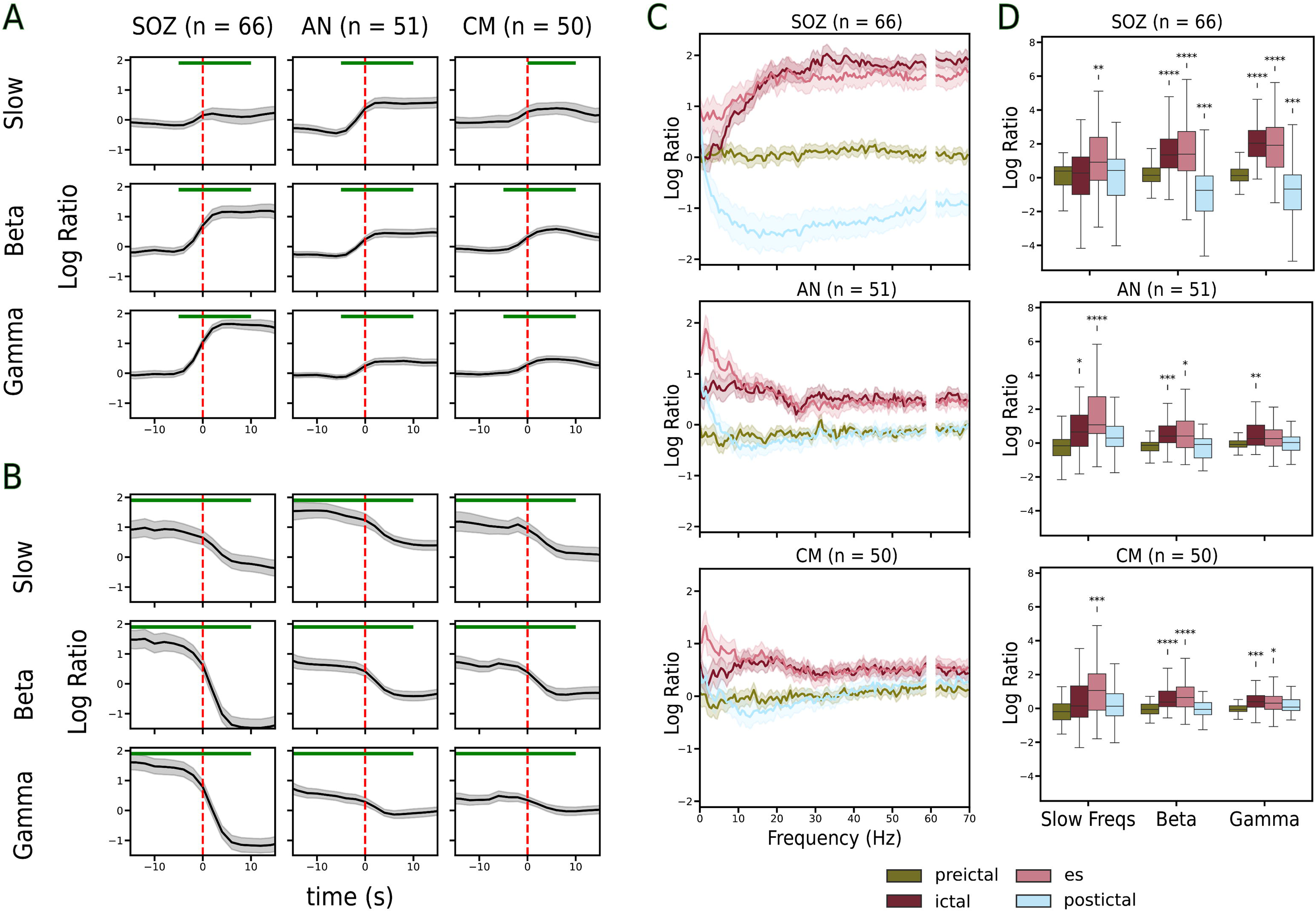
Frequency-specific spectral power dynamics in thalamus and seizure onset zone across seizure evolution. Spectral power was computed in canonical frequency bands (slow, 1–12 Hz; beta, 13–30 Hz; gamma, 30–70 Hz) and expressed as log-ratio relative to an interictal baseline. (A) Time-resolved spectral power changes aligned to seizure onset for the SOZ, ipsilateral AN (Ip AN), and ipsilateral CM (Ip CM), averaged across seizures. The vertical dashed line denotes seizure onset. (B) Same as (A), but time-aligned to seizure termination. The vertical dashed line denotes seizure termination. (C) Broadband spectral power changes in the SOZ, Ip AN, and Ip CM across seizure phases (preictal, ictal, end-of-seizure [ES], and postictal). (D) Box plots showing frequency-band–specific spectral power changes for each region and seizure phase. Statistical significance was assessed relative to the interictal baseline. *: 1.00e-02 < p <= 5.00e-02; **: 1.00e-03 < p <= 1.00e-02; ***: 1.00e-04 < p <= 1.00e-03; ****: p <= 1.00e-04

### Thalamocortical connectivity increased from seizure onset to termination

Undirected connectivity between each thalamic nucleus and the SOZ was quantified using iCoh. Time-series analyses demonstrated significant increases in thalamocortical iCoh during seizures, with elevations observed during both the ictal and ES periods, particularly in the slow and beta bands (**Fig. 3A,B**). Period-wise analyses further showed significant increases in thalamocortical slow- and beta-band iCoh during the ictal, ES, and postictal periods, whereas gamma-band iCoh increases were restricted to the ictal and ES periods (**Fig. 3C**).

**Figure 3.**
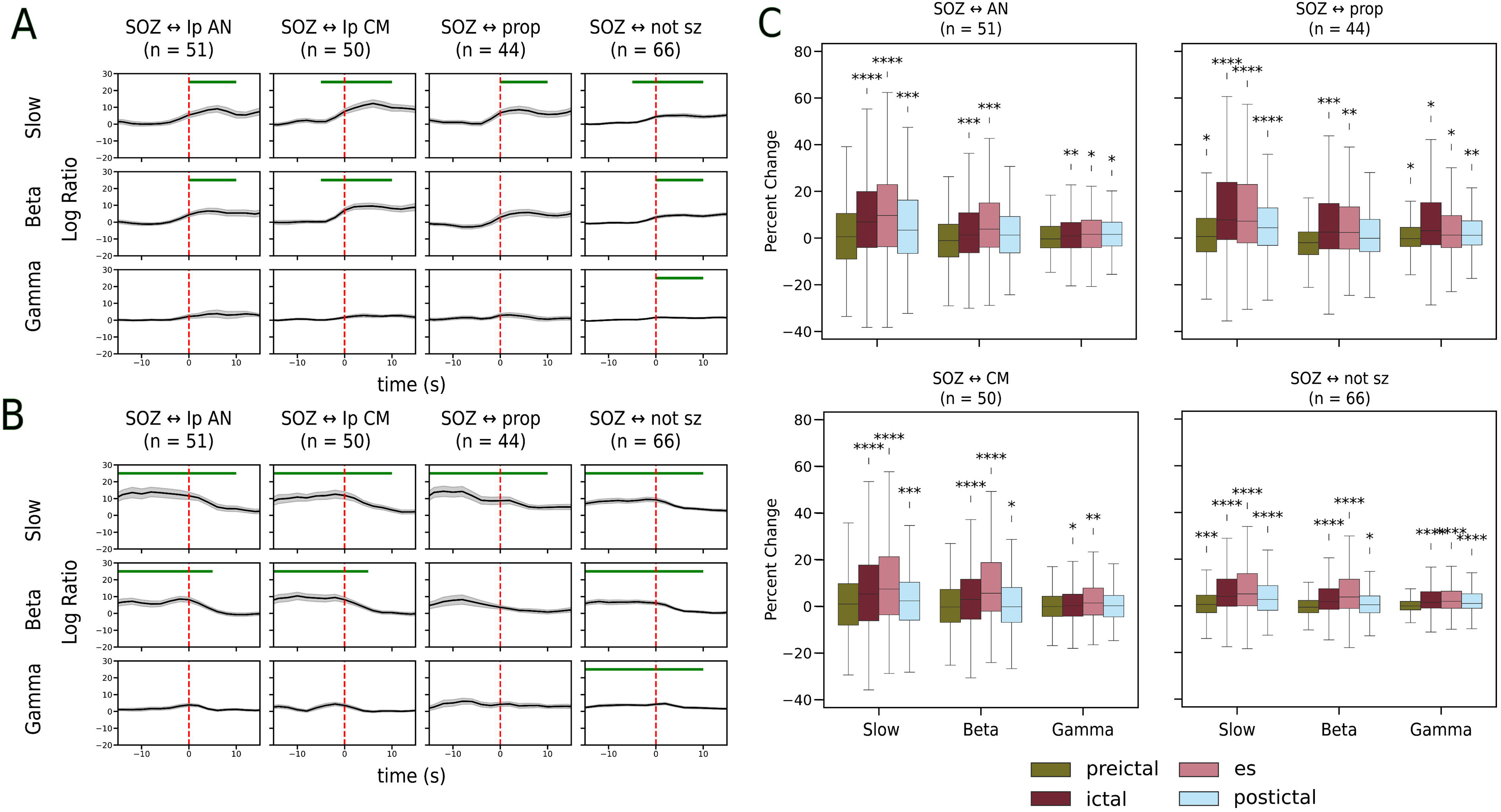
Thalamocortical and cortico-cortical connectivity dynamics assessed using imaginary coherence. Imaginary coherence (iCoh) was computed between the SOZ and ipsilateral thalamic nuclei (Ip AN and Ip CM), as well as between the SOZ and cortical regions (propagation and non-seizure areas), in canonical frequency bands and expressed as percent change relative to an interictal baseline. (A) Time-resolved iCoh changes aligned to seizure onset for thalamocortical (AN–SOZ, CM–SOZ) and cortico-cortical (SOZ–propagation, SOZ–non-seizure) interactions, averaged across seizures. The vertical dashed line denotes seizure onset. Horizontal bars indicate time periods with statistically significant increases relative to baseline. (B) Same as (A), but time-aligned to seizure termination. The vertical dashed line denotes seizure termination. Horizontal bars indicate time periods with statistically significant increases relative to baseline. (C) Box plots showing frequency-specific iCoh changes for thalamocortical and cortico-cortical interactions across seizure phases (preictal, ictal, end-of-seizure [ES], and postictal). Statistical significance was assessed relative to the interictal baseline. *: 1.00e-02 < p <= 5.00e-02; **: 1.00e-03 < p <= 1.00e-02; ***: 1.00e-04 < p <= 1.00e-03; ****: p <= 1.00e-04

Similarly, directed connectivity assessed using GCA demonstrated bidirectional increases between the thalamic nuclei and the SOZ during both the ictal and ES periods, again most prominently in the slow and beta bands (**Fig. 4A,B**). Box-plot analyses confirmed significant bidirectional increases relative to the interictal baseline (**Fig. 4C**). During the ictal period, no significant difference was observed between inflow and outflow in the AN for either the slow band (p = 0.052) or beta band (p = 0.759). Similarly, CM slow-band connectivity showed no significant inflow–outflow difference (p = 0.205), whereas CM beta-band connectivity exhibited a significant asymmetry between inflow and outflow (p = 0.000325). At the ES period, no significant differences between inflow and outflow were observed in either frequency band for AN (slow: p = 0.0822; beta: p = 0.712) or CM (slow: p = 0.628; beta: p = 0.188), indicating balanced bidirectional thalamocortical coupling toward seizure termination.

**Figure 4.**
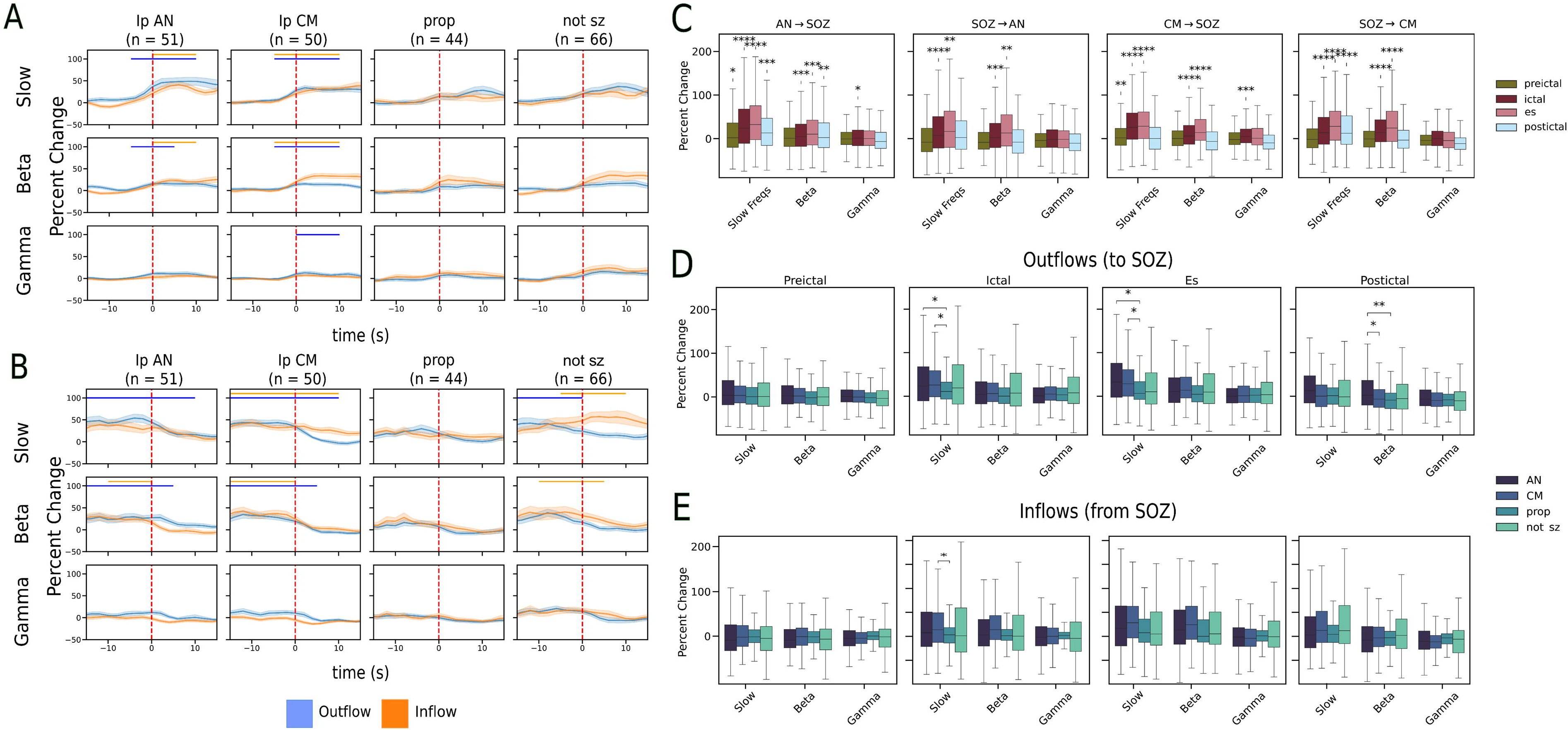
Directed thalamocortical and cortico-cortical connectivity dynamics assessed using spectral Granger causality. Spectral Granger causality (GCA) was computed to quantify directed connectivity between the SOZ and ipsilateral thalamic nuclei (Ip AN and Ip CM), as well as between the SOZ and cortical regions (propagation and other non-seizure cortical regions), in canonical frequency bands and expressed as percent change relative to an interictal baseline. Directed connectivity was separated into inflow (SOZ → target region) and outflow (target region → SOZ). (A) Time-resolved spectral GCA aligned to seizure onset for both thalamocortical (AN–SOZ, CM–SOZ) and cortico-cortical (SOZ–propagation, SOZ–other cortical) interactions, showing inflow and outflow. The vertical dashed line denotes seizure onset. Horizontal bars indicate time periods with statistically significant increases relative to baseline. (B) Same as (A), but time-aligned to seizure termination. The vertical dashed line denotes seizure termination. Horizontal bars indicate time periods with statistically significant increases relative to baseline. (C) Box plots showing frequency-specific spectral GCA changes for thalamocortical and cortico-cortical inflow and outflow across seizure phases (preictal, ictal, end-of-seizure [ES], and postictal), relative to the interictal baseline. (D) Box plots comparing slow-band outflow directed toward the SOZ from thalamic nuclei (AN and CM) and from cortical regions (propagation and other cortical regions) across seizure phases. (E) Box plots comparing slow-band inflow directed from the SOZ to thalamic nuclei (AN and CM) and to cortical regions (propagation and other cortical regions) across seizure phases. Statistical significance was assessed relative to the interictal baseline or between network types, as appropriate. *: 1.00e-02 < p <= 5.00e-02; **: 1.00e-03 < p <= 1.00e-02; ***: 1.00e-04 < p <= 1.00e-03; ****: p <= 1.00e-04

### Comparison between thalamocortical and cortico-cortical ictal dynamics

We next examined whether the magnitude of directed network activation differed between thalamocortical networks (AN–SOZ and CM–SOZ) and cortico-cortical networks (SOZ–propagation zone). Slow-band outflow toward the SOZ was significantly greater in thalamocortical networks than in cortico-cortical networks during both the ictal and ES periods (**Fig. 4D,E**). Specifically, slow-band outflow from AN to the SOZ exceeded outflow from propagation zones to the SOZ during the ictal period (p = 9.06 × 10L^3^) and ES period (p = 8.80 × 10L^3^). Similarly, slow-band outflow from CM to the SOZ exceeded cortico-cortical outflow during the ictal period (p = 3.30 × 10L^3^) and ES period (p = 5.7 × 10L^3^). Moreover, slow-band inflow from the SOZ to the CM was significantly greater than cortico-cortical inflow from the SOZ to propagation regions. No significant differences were observed between thalamocortical and SOZ–other cortical networks across frequency bands.

To contextualize thalamocortical effects, power and iCoh were also quantified in propagation and non-seizure cortical regions. Spectral power in propagation and non-seizure regions increased from seizure onset to termination, but to a lesser magnitude than in the SOZ and thalamus (**Supplementary Figure 1**). Similarly, coherence increased in both thalamocortical and cortico-cortical networks, including SOZ–propagation and SOZ–non-seizure interactions (**Fig. 3C**).

### Thalamocortical connectivity varies by seizure focus and onset patterns

We next examined whether thalamocortical connectivity differed according to seizure focus, ictal onset pattern, and presence of diffuse cortical involvement at seizure onset.

#### Seizure focus

When stratified by seizure focus, significant increases in AN–SOZ connectivity were observed in the slow and beta bands during the ictal and ES periods in limbic seizures (**Fig. 5A**). More modest increases were observed in frontal seizures, primarily in the slow band during ES, whereas temporal seizures showed increased beta-band connectivity during the ictal period. In the CM–SOZ network, connectivity increased across all seizure foci, with statistically significant increases in limbic and parietal seizures (**Fig. 5A**).

**Figure 5.**
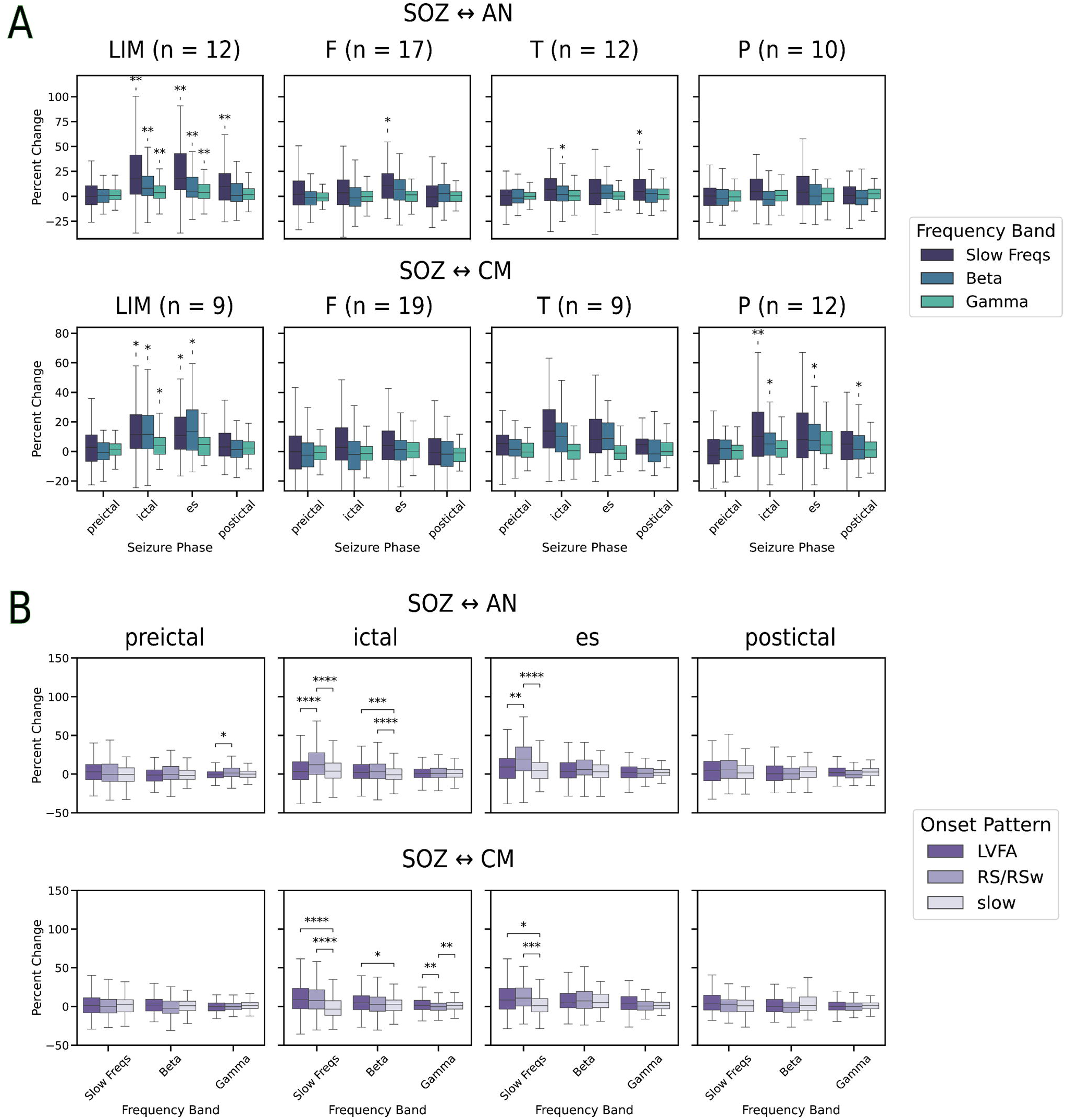
Subgroup analysis of thalamocortical connectivity using imaginary coherence. Imaginary coherence (iCoh) was computed between the SOZ and ipsilateral thalamic nuclei (Ip AN and Ip CM) in canonical frequency bands and expressed as percent change relative to an interictal baseline. (A) Seizure focus. Frequency-specific iCoh changes for AN–SOZ (top) and CM–SOZ (bottom) interactions stratified by seizure focus (frontal, parietal, temporal, occipital, and limbic) across seizure phases (ictal and end-of-seizure [ES]). (B) Ictal onset pattern. Frequency-specific iCoh changes for AN–SOZ (top) and CM–SOZ (bottom) interactions stratified by ictal onset pattern (LVFA, RS/RSw, and other) across seizure phases. Statistical significance was assessed relative to the interictal baseline. *: 1.00e-02 < p <= 5.00e-02; **: 1.00e-03 < p <= 1.00e-02; ***: 1.00e-04 < p <= 1.00e-03; ****: p <= 1.00e-04

#### Ictal onset pattern

When seizures were stratified by ictal onset pattern, RS/RSw patterns demonstrated significantly greater thalamocortical slow-band connectivity increases than LVFA and other patterns in the AN–SOZ network (**Fig. 5B**). In the beta band, both LVFA and RS/RSw patterns showed greater connectivity increases than other patterns in the AN–SOZ network. In the CM–SOZ network, slow-band connectivity increased significantly in both LVFA and RS/RSw patterns compared with other onset patterns (**Fig. 5B**).

#### Diffuse cortical involvement at onset

Finally, we examined whether diffuse cortical involvement at seizure onset influenced thalamocortical connectivity. We found that seizures with diffuse onset exhibited lower thalamocortical activity in the ictal period across the frequency bands (**Supplementary Figure 2**).

### Seizure state can be identified from thalamocortical features

We next evaluated whether thalamocortical features could be used to classify seizure state using random forest classifiers with leave-one-patient-out cross-validation (**Fig. 6A**). Mean ± standard deviation AUROC values across patients were reported to quantify classification performance. To avoid extreme cases that could bias model evaluation, very short (<10 s) and very long (>300 s) seizures were excluded from this analysis (n = 15 seizures excluded). For the AN-based model, spectral features alone achieved an AUROC of 0.785 ± 0.182, which increased to 0.803 ± 0.176 with the addition of iCoh and to 0.817 ± 0.168 with the inclusion of GCA. The full feature set (spectral power + iCoh + GCA) yielded the highest AN performance, with an AUROC of 0.825 ± 0.163 (**Fig. 6B**). A similar performance pattern was observed for the CM-based model. Spectral features alone achieved an AUROC of 0.777 ± 0.176, increasing to 0.793 ± 0.171 with iCoh and to 0.840 ± 0.147 with GCA. The full CM feature set produced the highest performance, with an AUROC of 0.839 ± 0.149 (**Fig. 6C**).

**Figure 6.**
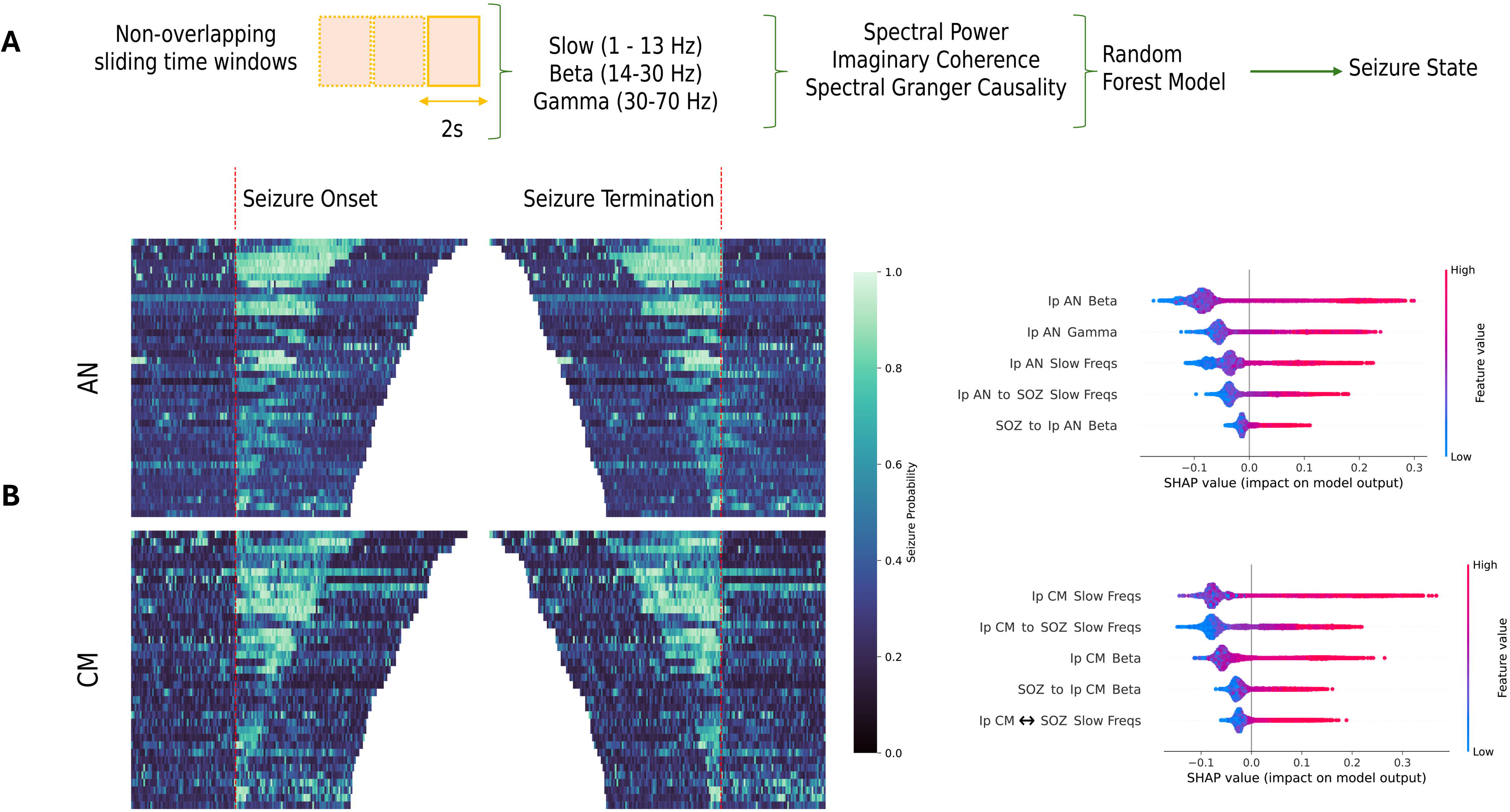
Seizure state identification using thalamic spectral power and thalamocortical connectivity features. (A) Overview of the seizure state prediction pipeline. Each seizure was segmented into non-overlapping two-second windows spanning one minute before seizure onset to one minute after seizure termination. For each window, thalamic spectral power, iCoh, and GCA between the thalamus and SOZ were computed. Random Forest classifiers were trained to classify ictal versus non-ictal states using leave-one-patient-out cross-validation. (B) AN-based classifier performance and feature importance. Left: Heatmaps showing predicted seizure probabilities for each two-second window using AN features, ordered by seizure duration. Data are aligned to seizure onset (left panel) and seizure termination (right panel). Right: SHAP summary plot showing feature importance for the AN classifier. Features are ordered by mean absolute SHAP value; the x-axis indicates the direction and magnitude of each feature’s contribution to ictal classification (positive values indicate higher ictal probability), and color denotes feature magnitude (red, high; blue, low). (C) CM-based classifier performance and feature importance. Same format as (B), but using CM features. Heatmaps display predicted seizure probabilities aligned to seizure onset and termination, and SHAP summary plots illustrate feature importance for the CM classifier.

To identify the most informative features contributing to seizure state classification, SHAP values were computed and averaged across cross-validation folds. For the AN model, the most predictive features included broadband AN spectral power (slow, beta, and gamma bands), slow-band outflow from the AN to the SOZ, and beta-band inflow from the SOZ to the AN. For the CM model, the strongest predictors included slow- and beta-band CM spectral power, slow-band outflow from the CM to the SOZ, beta-band inflow from the SOZ to the CM, and bidirectional slow-band iCoh.

## Discussion

Across 19 patients with pediatric-onset DRE and 66 focal seizures, we show that seizures recruit a robust, frequency-specific thalamocortical network that evolves systematically from onset through termination. Thalamic involvement was common at seizure onset and became near-universal by seizure termination, with a stereotyped evolution in visual patterns from LVFA at onset to more RS/RSw near the ES. Quantitatively, the thalamus and SOZ exhibited prominent early broadband power increases that attenuated toward termination, while thalamocortical connectivity (iCoh and GCA) increased most strongly in the slow and beta bands across the seizure course, reaching maximal expression around the ES epoch and then rapidly diminishing at termination. Directed analyses demonstrated bidirectional thalamocortical coupling, with slow-frequency thalamus→SOZ outflow emerging as a distinctive feature that exceeded propagation-zone→SOZ outflow. Finally, seizure state was classifiable using thalamic spectral power and thalamocortical connectivity features, supporting a mechanistic and actionable framework for the network dynamics (**Figure 7**).

**Figure 7.**
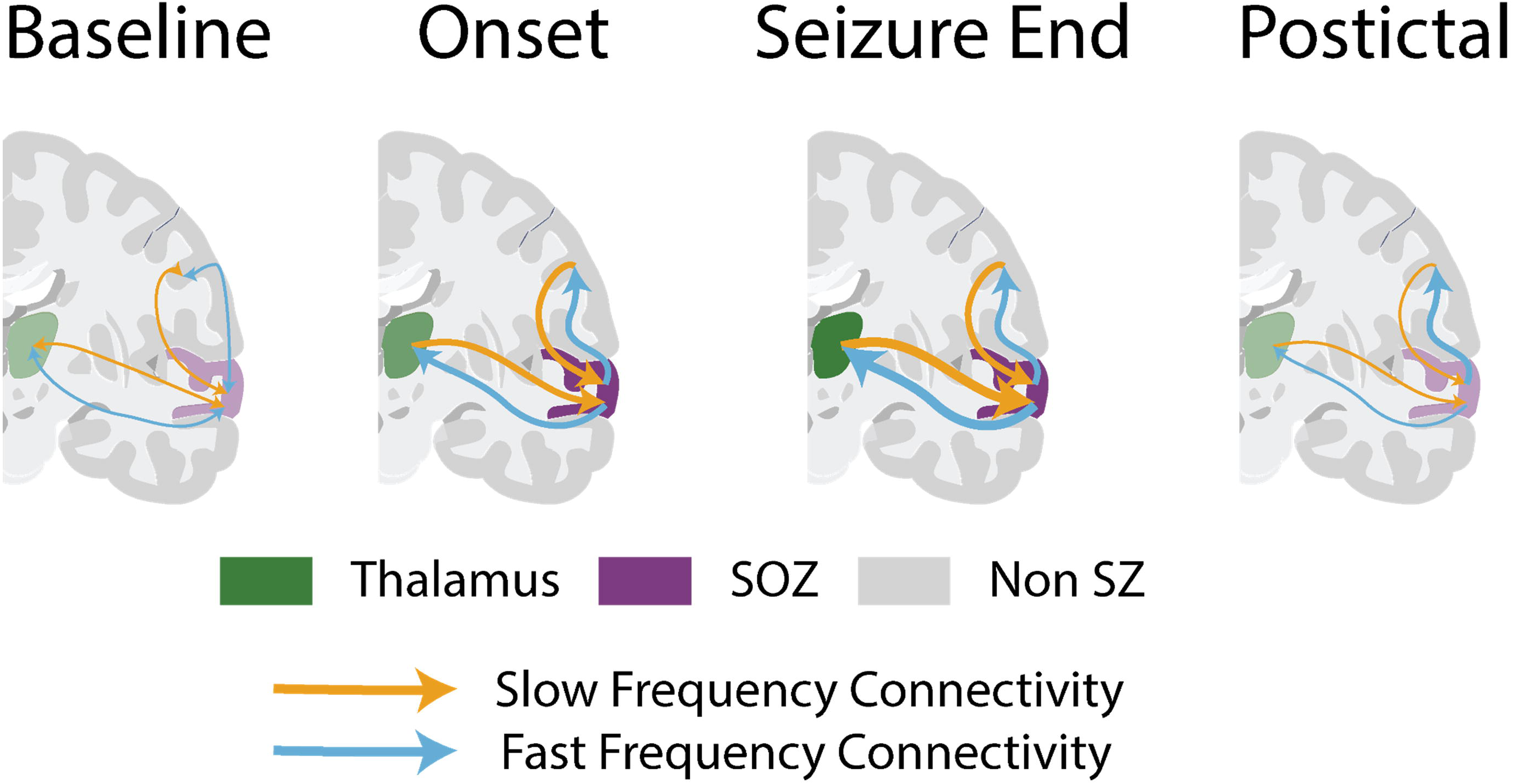
Hypothesized model of frequency-specific thalamocortical network dynamics during focal seizures. Schematic illustration summarizing a hypothesized model of frequency-specific thalamocortical and cortico-cortical network dynamics across seizure evolution. At seizure onset, cortical regions within the SOZ exhibit prominent fast-frequency activity that is primarily organized around the SOZ, with concurrent engagement of thalamocortical networks. As seizures progress toward the end-of-seizure (ES) period, slow-and beta-band thalamocortical connectivity—particularly slow-frequency outflow from the thalamus (AN and CM) to the SOZ—reaches maximal expression, with bidirectional thalamocortical interactions maintained during this late ictal phase. At seizure termination, this organized thalamocortical coupling rapidly collapses, consistent with a loss of frequency-specific thalamocortical network engagement rather than gradual attenuation. Findings from seizure state classification models converge with this framework: ictal states are characterized by the presence of slow-frequency thalamus-to-SOZ outflow together with fast-frequency inflow from the SOZ to the thalamus, whereas the absence of this bidirectional, frequency-specific signature corresponds to non-ictal states, including the post-termination period. Cortico-cortical interactions are also engaged throughout seizures but exhibit comparatively weaker, consistent with an ictal cortical network that remains primarily organized around the SOZ. Collectively, this conceptual model highlights frequency- and direction-specific thalamocortical interactions—and their abrupt disengagement at seizure termination—as potential targets for adaptive neuromodulation strategies.

### Clinical and translational implications for personalized thalamic neuromodulation

Our results directly inform thalamic neuromodulation strategies that are already in broad use for patients who are not candidates for curative resection. The efficacy of DBS targeting the AN in focal epilepsy has been established,^2^ and additional thalamic stimulation strategies have been explored, including CM targeting in generalized and multifocal phenotypes such as Lennox–Gastaut syndrome and pediatric-onset epilepsy.^4–6^ Our data add a translationally important layer: the thalamus provides frequency- and direction-specific signatures that are measurable in real time and generalize across patients to support seizure-state detection, suggesting that adaptive stimulation could be triggered not simply by local power changes but by specific thalamocortical interaction motifs. This aligns with the broader push toward network-guided neuromodulation^33^ and with emerging hodology-driven personalization of stimulation targets and parameters.^34^ In addition, our nucleus- and syndrome-dependent findings—such as stronger AN–SOZ interactions in limbic seizures—support leveraging known anatomical connectivity when selecting thalamic targets and patient-specific stimulation programs.

### Thalamocortical network involvement as a core feature of focal seizures

The concept that seizures engage thalamocortical loops has deep historical roots, from classic “centrencephalic” frameworks proposed by Penfield to modern mechanistic views in which pathological activity hijacks physiological thalamocortical circuitry.^19, 21, 35^ Animal studies of absence epilepsy provided early causal insights, demonstrating that a cortical focus can drive widespread corticothalamic networks.^36^ In focal epilepsies, a growing human SEEG literature has documented early thalamic recruitment and thalamocortical coupling,^7–9, 14^ including evidence that thalamic recruitment can vary across seizures within individuals.^13^ Our work extends these foundations by explicitly tracking frequency-resolved thalamocortical dynamics from onset through termination, directly comparing them with cortico-cortical dynamics, and demonstrating that thalamic spectral and network signatures are sufficiently informative to support seizure-state decoding. These findings are consistent with emerging concepts that the thalamus can exert frequency-coded control over telencephalic ictal organization.^37^

### Relationship between ictal thalamocortical and cortico-cortical networks

Cortico-cortical network dynamics remain a core feature of focal seizures, and our findings are consistent with prior reports describing increased synchronization and structured propagation across cortical networks.^15–18^ Importantly, our data support a nuanced relationship between cortical and thalamocortical dynamics: both networks are engaged, but slow-frequency outflow toward the SOZ was comparatively stronger in thalamocortical networks than in propagation-zone→SOZ cortico-cortical interactions. This is consistent with an ictal cortico-cortical network that is primarily organized around the SOZ, with limited engagement of long-range synchronized cortical interactions. This observation is relevant to interictal network frameworks such as the interictal suppression hypothesis, which proposes that non-SOZ regions can suppress SOZ activity,^38, 39^ as well as to recent work further evaluating suppression-related network signatures.^40^ Seizure onset may therefore involve a transient breakdown of these interictal network structures, such that ictal cortico-cortical interactions predominantly reflect SOZ-driven propagation rather than effective inhibitory feedback toward the SOZ. Visual EEG analysis patterns further support state dependence: LVFA was common at onset, a pattern associated with reduced synchrony and local microcircuit changes early in seizures,^41, 42^ whereas RS/RSw patterns were more prominent near ES, aligning with the emergence of synchronized slow/beta thalamocortical engagement that may be particularly relevant to seizure termination (**Figure 5**). Consistent with this interpretation, LVFA-onset and diffuse-onset seizures showed relatively reduced thalamocortical engagement in specific frequency bands during the ictal period, whereas RS/RSw-associated connectivity increases were more prominent later in the seizure course (**Supplementary Figure 2**).

### Mechanistic considerations and implications for seizure termination

Corticothalamic inputs shape thalamocortical network activity, supporting physiological rhythms such as sleep spindles^35^ or, under pathological conditions, facilitating epileptic synchronization.^43^ It remains unresolved whether the thalamus primarily amplifies seizures, restrains them, or serves both roles in a state-dependent manner; however, experimental and clinical evidence supports this dual functional role. Closed-loop optogenetic inhibition of the thalamus was shown to interrupt seizures in a rat model of post-cortical injury epilepsy, where thalamocortical neurons connected to the injured cortex became hyperexcitable and necessary for seizure maintenance.^44^ Complementary animal work further demonstrates that the thalamus can exert frequency-dependent control over large-scale coherence to actively initiate or maintain ictal states, and that pharmacological thalamic inhibition disrupts ongoing seizures.^37^ In this context, our data are compatible with a rapid-disengagement model of termination,^20^ in which seizure termination reflects not gradual exhaustion but a sudden loss of organized, frequency-specific coupling that sustains the ictal network (**Figure 7**). Such abrupt ictal network disconnection may reflect active recruitment of inhibitory thalamic circuitry, particularly thalamic reticular nucleus–mediated suppression of thalamocortical relay output, which has been shown to rapidly abort ongoing seizures when engaged. ^45, 46^ Together, these findings support a state-dependent model in which early ictal recruitment of the thalamus is driven by excitatory glutamatergic corticothalamic input, whereas late-phase seizure termination may be mediated by engagement of GABAergic thalamic inhibitory networks that suppress thalamocortical coupling.

This framework aligns with stimulation studies emphasizing network-level mechanisms of therapeutic benefit.^47^ Clinically, CM-targeted DBS in Lennox–Gastaut syndrome further supports the relevance of thalamocortical network targeting and connectivity-informed programming.^48^ Together, these findings motivate a testable translational hypothesis: adaptive or closed-loop thalamic neuromodulation could detect thalamic power changes or the emergence of a frequency- and direction-specific thalamocortical signature, such as slow-band thalamus→SOZ outflow coupled with fast- or beta-band SOZ → thalamus inflow, and deliver stimulation either early, to prevent stabilization of the ictal network, or late, to facilitate thalamocortical decoupling and promote earlier termination, with parameters guided by patient-specific hodology.^33, 34^

## Limitations

Several limitations should be considered. The cohort size was modest and clinically heterogeneous, with anatomically diverse seizure onset zones, limiting fine-grained subgroup inference despite enhancing generalizability. Thalamic sampling was restricted to AN and CM and was clinically determined, limiting inference about other nuclei such as the pulvinar. Connectivity measures such as iCoh and spectral GCA are informative but indirect and can be influenced by common drivers, volume conduction, and nonstationarity. Finally, seizure termination likely reflects the interaction of multiple processes, and the retrospective nature of this study limits our ability to determine whether any single network feature plays a primary mechanistic role.

## Future perspectives

Future studies should prioritize larger, multicenter cohorts with broader thalamic and cortical sampling, including additional nuclei implicated in focal seizure propagation and termination, and should integrate structural and functional imaging to relate iEEG dynamics to individualized connectomes. Prospective work is needed to test whether frequency- and direction-specific thalamocortical signatures improve adaptive or closed-loop stimulation relative to conventional power-based detectors, and whether targeting late-ictal thalamocortical coupling can reliably accelerate termination without disrupting physiological thalamocortical functions.

## Supplementary material

Supplementary material is available at *Brain* online.

