## Supplemental document for "Distinct Spectral and Directional Thalamocortical Network Dynamics Define Focal Seizure Evolution"

**Supplementary documents**

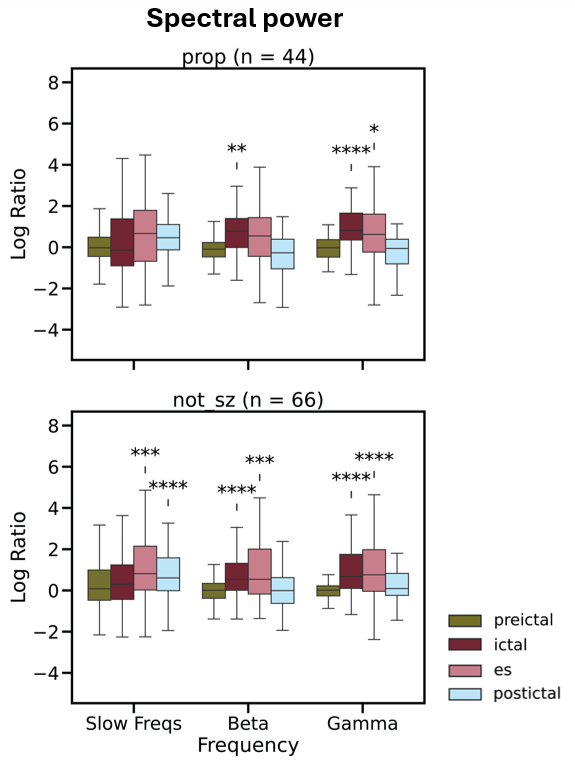

**Supplementary Figure 1. Frequency-specific spectral power dynamics in non-seizure onset zone cortical regions across seizure evolution.** Box plots showing frequency-band–specific spectral power changes for each region and seizure phase. Statistical significance was assessed relative to the interictal baseline.

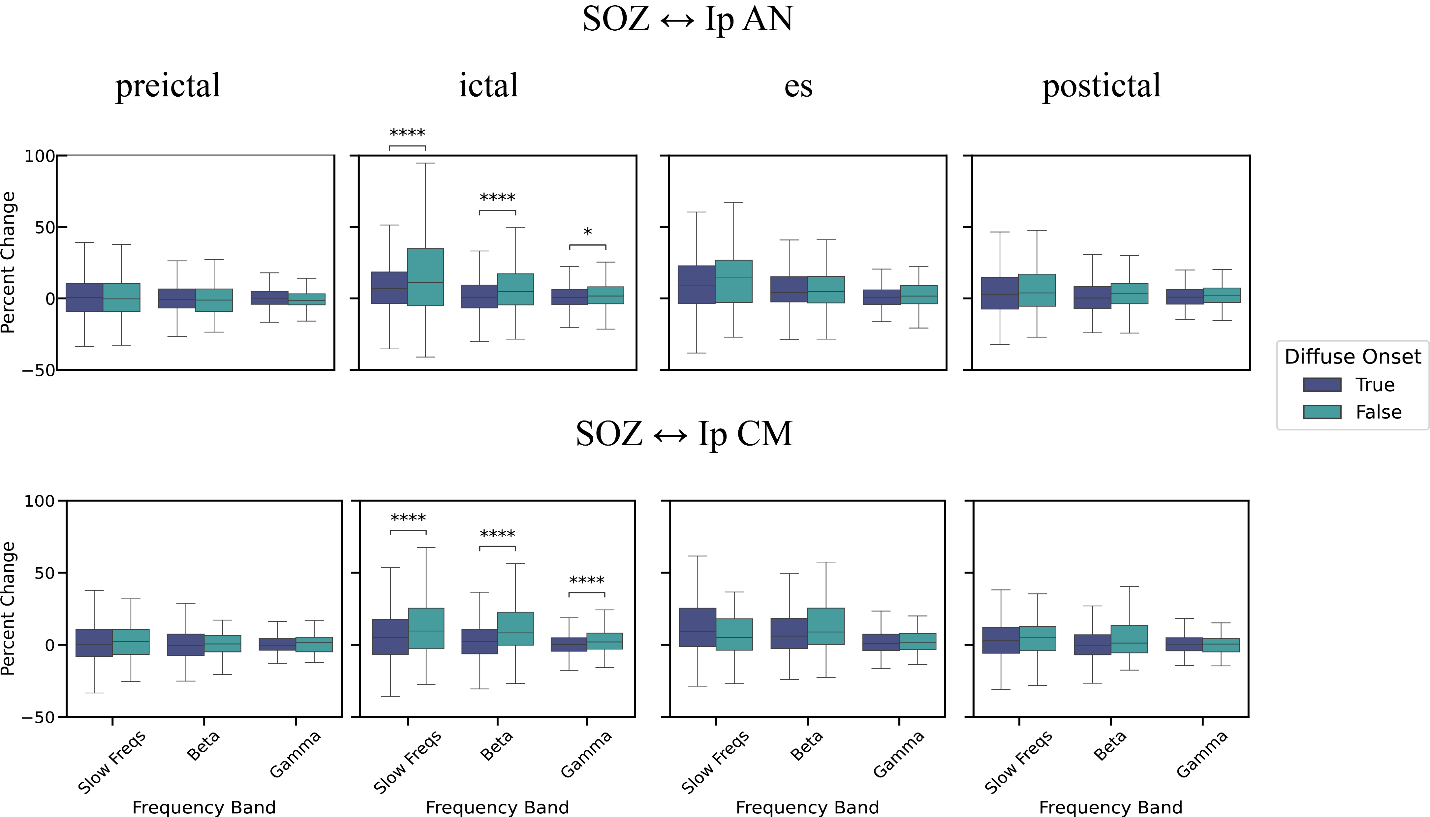

**Supplementary Figure 2. Subgroup analysis based on onset pattern for thalamocortical connectivity dynamics across seizure phases.** Imaginary coherence (iCoh) was computed between the SOZ and ipsilateral thalamic nuclei (Ip AN and Ip CM) in canonical frequency bands and expressed as percent change relative to an interictal baseline. Frequency-specific iCoh changes for AN–SOZ (top) and CM–SOZ (bottom) interactions stratified by seizure onset pattern (diffuse vs. not diffuse).
